# Prevalence, risk factors and adverse outcomes for intrapartum related asphyxia amongst newborns from mothers with low-risk deliveries in peri-urban Gambian health facilities: a retrospective cohort study

**DOI:** 10.1101/2025.10.30.25339034

**Authors:** Nathalie Beloum, Bully Camara, Usman N. Nakakana, Fatoumata Sillah, Madikoi Danso, Joquina C. Jones, Shashu Graves, Edrissa Sabally, Siaka Badjie, Omar Jarra, Abdoulie Suso, Ebrahim Ndure, Yusupha Njie, Christian Bottomley, Halidou Tinto, Umberto D’Alessandro, Helen Brotherton, Anna Roca

**Author notes:** **Corresponding author & contact details:** Dr Nathalie Beloum.

## Abstract

**Introduction:** Despite significant progress in perinatal care, intrapartum related asphyxia remains a leading cause of neonatal morbidity and mortality, especially in the poorest regions of the world such as sub-Saharan Africa. The objective of this study is to determine the prevalence, risk factors, and adverse outcomes of intrapartum related asphyxia among low risk health facility based deliveries in The Gambia.

**Methods:** This is a secondary analysis of a randomised clinical trial conducted in western Gambia between October 2017 and May 2021. Women with low risk pregnancies in labour and their live neonates were recruited at two public health facilities. The prevalence, risk factors, and neonatal outcomes of intrapartum related asphyxia, defined as 1 minute Apgar score <7, were determined using descriptive methods, and unadjusted and adjusted logistic regression.

**Results:** Among 6,758 neonates, 257 (3.8%) experienced intrapartum related asphyxia, 93 (36.2%) of them required hospitalisation. The neonatal case fatality rate was 13.9% (34/245) among newborns with intrapartum related asphyxia. Neonatal death (OR=19.56, 95% CI 12.02 to 31.55, p value<0.001) and hospitalisation (OR=11.91, 95% CI 8.90 to 15.90, p value<0.001) were significantly higher among these neonates. Severe intrapartum related asphyxia cases had a significantly high risk of neonatal death (aOR:71.8, 95% CI 36.00 to 140.40, p value<0.001) and hospitalisation (aOR:19.5, 95% CI 10.80 to 35.00, p value<0.001); however, this risk remained considerable even in mild to moderate cases. Adjusted analysis revealed macrosomia as a major modifiable risk factor (aOR: 3.31, 95% CI 1.78 to 6.15, p value<0.001), and several identifiable risk factors such as history of previous stillbirths (aOR: 2.79, 95% CI 1.41to 5.52, p value=0.003) or miscarriage (aOR: 1.65, 95% CI 1.08 to 2.52, p value=0.02),and primiparity (aOR: 1.69, 95% CI 1.16 to 2.46, p value<0.001).

**Conclusions:** Intrapartum related asphyxia is a major contributor to neonatal morbidity and mortality among low risk pregnant women in peri urban Gambia. A significant proportion of intrapartum related asphyxia is preventable with timely identification of risk factors by improved antenatal and obstetric care followed by appropriate intrapartum management. Availability of adequate equipment, infrastructures, and skilled staff to help neonates breathe at birth are urgently required to reach the Sustainable Development Goal 3.2 of reducing by 2030 neonatal mortality to 12 deaths per 1000 live births.

**Trial Registration:** NCT03199547: Clinicaltrials.gov. Registered on 23rd June 2017

## Background

In 2021, 2.3 million neonates died worldwide, representing approximately half of all under-five deaths (1). Intrapartum-related asphyxia (IRA), also known as *birth asphyxia* remains a leading cause of neonatal mortality, contributing to 24% of neonatal deaths worldwide (1). In addition, IRA leads to short- and long-term morbidities, including early hospitalisation, neurodevelopment and motor delays, seizures, or cerebral palsy (2). According to the World Health Organization, in 2019 IRA accounted for 61 million disability-adjusted life years (DALYs) (3). In sub-Saharan Africa, the region with the highest burden of neonatal mortality, IRA was the leading cause (30%) of overall neonatal deaths by day 28 in 2019, 60% of which occurred during the first week of life (1).

The main risk factors associated with IRA can be categorised as maternal, pregnancy-related, or neonatal. Maternal factors include socio-demographic characteristics such as age, educational status, and marital status (4). In addition, pregnancy-related conditions such as primiparity, severe maternal hypotension or hypertension/pre-eclampsia, and antepartum haemorrhage (5) are important in the causal pathway for IRA. Labour complications such as placental abruption or umbilical cord prolapse are major intrapartum sentinel events leading to IRA, with meconium-stained amniotic fluid a physical manifestation of foetal distress (6). In sub-Saharan Africa, additional neonatal risk factors such as birth weight (low or macrosomia), multiple gestations, and preterm delivery (7, 8) have been previously identified.

IRA can be identified by three different types of measurements: (i) process-based indicators (i.e., prolonged labour or foetal distress detected via partograph or cardiotocography); (ii) clinical sign-based indicators (i.e., low Apgar scores, foetal or neonatal acidosis), and (iii) outcome-based indicators (i.e., stillbirth or neonatal mortality, signs of neonatal encephalopathy) (9). In sub-Saharan Africa and other low-income regions, the Apgar score remains routinely and widely used to detect IRA. The Apgar score is a quick assessment performed on a neonate after birth and comprises the following five clinical signs: heart rate, respiratory effort, muscle tone, reflex irritability, and colour. Each sign is assigned a value of 0, 1, or 2, with the sum comprising the score out of 10 (10). The Apgar score is reported at 1-minute and 5-minutes after birth for all neonates, and at 5-minute intervals thereafter until 20 minutes for infants with a score less than 7 (10). A 1-minute Apgar score provides an indication of intrapartum health and the neonatal response to the process of birth (11, 12). However, Apgar scores can be subjective in the absence of robust training and prone to intra- and inter-observer variability (13). With limited training, values of Apgar scores are often over-estimated thus underestimating the burden of IRA in resource-constrained settings as only the more severe cases are identified. Despite these limitations, understanding the factors associated with a low 1-min Apgar score may help to improve the transition from intrauterine to extrauterine life (14), enable life-saving resuscitation for those at highest risk, and eventually decrease the mid and long-term sequelae from IRA.

Evidence on risk factors for IRA in West Africa is limited, despite its contribution towards adverse neonatal outcomes in this region of high neonatal mortality. In the Gambia, a small country in the West African region, the neonatal mortality rate (NMR) in 2020 was estimated at 26 deaths per 1000 live births (15), more than 2-fold higher than the target of 12 deaths per 1000 live births set in the Sustainable Development Goal (SDG) for 2030. In a cohort study that included 829 deliveries at a Gambian urban health facility, IRA accounted for 31% of neonatal deaths, representing the second most common diagnosis (16).

The study presented here aims to determine the prevalence and the risk factors associated with IRA as detected by the 1-min Apgar score, in a large cohort of newborns at two Gambian peri-urban health facilities. Detailed and context-specific understanding of the risk factors associated with IRA has the potential to improve perinatal care and optimise the transition to extra-uterine life (17).

## Methods

### Study design

This is a secondary analysis of the PregnAnZI-2 trial (18) a phase III, double-blind, placebo-controlled individually randomised clinical trial in which *∽*12,000 women in labour in The Gambia and Burkina Faso received either azithromycin or placebo (ratio 1:1) from October 2017 to May 2021 (18). The trial showed no differences in IRA by trial arm (19) and therefore we have included the full Gambian cohort in this retrospective cohort study. A separate analysis is conducted for the Burkina Faso cohort.

### Study setting

The climate of The Gambia is typical of the sub-Sahel region, with a long dry season from November to May and a short rainy season between June and October. The population of the catchment area is representative of The Gambia and includes all the main ethnic groups. Illiteracy rates are high, up to 55% among women (18).

The study was conducted at two Government-run health facilities situated in western Gambia, namely Serekunda Health Centre (SHC) and Bundung Maternal and Child Health Hospital (BMCHH) (18). These two health facilities could not consistently provide obstetric ultrasound scans during antenatal visits throughout our study period. At the time of data collection, SHC could not provide emergency obstetric care, including surgical care; and lacked a neonatology unit. Thus, women in need of an emergency caesarean section were referred to the Kanifing General Hospital, 3.4km away from SHC. Neonates born at SHC and requiring hospital care were transferred to the neonatal unit at Kanifing General Hospital or to BMCHH. BMCHH provided emergency obstetric and neonatal care and on rare occasions transferred women or neonates to the national referral centre, the Edward Francis Small Teaching Hospital in Banjul, about 15.5 km from BMCHH.

### Participants

The PregnAnZI-2 trial recruited pregnant women aged >16 years receiving antenatal, and intrapartum care at the study health facilities. Women were excluded if they had known HIV infection, any chronic or acute conditions, planned (elective) caesarean section or known required referral, known severe congenital malformation (detected antenatally), intrauterine death or allergy to macrolides, and drugs known to prolong QT interval taken during the last 2 weeks, such as chloroquine, quinine, piperaquine, and erythromycin (18). All women recruited in The Gambia who delivered a live-born neonate at a trial health facility were included in this secondary analysis.

### Study procedures

Written informed consent to participate in the trial was taken during antenatal visits and verbally confirmed during labour. As per trial procedures, socio-demographic and perinatal clinical data were transcribed from the participants’ antenatal card and partograph (the timing of labour was not recorded in our study) to an electronic case report form (eCRF) before discharge by trained nurses (18). This included the Apgar scores, and the status of the neonate at birth (alive or stillbirth) which were assigned contemporaneously by health facility nurses, midwives, or doctors. The delivery room health workers were trained on Apgar scoring relevance and methodology by research personnel shortly after the recruitment commenced, aiming to reduce potential subjectivity in scoring. Stillbirth was determined according to physical appearance, absence of heartbeat, and respiratory effort.

There was both active and passive surveillance by the trial team to collect information on the health status and evaluate the safety of the intervention for mothers and neonates (18). A thorough clinical assessment of the mother and the neonates, including the neonate’s survival status at the time of assessment (alive or dead) was carried out by either a trained trial research clinician or a paediatrician 4–24h after delivery (18) and before discharge. A further assessment was conducted at day 28 post-delivery, when mothers and their neonates were visited at home by trained trial nurses. Neonates requiring further examination by the trial clinicians were referred to the trial sites. There was passive follow-up of adverse events throughout the neonatal period, with mothers and neonates advised to visit the trial sites if they had health concerns. The trial clinician or a paediatrician then assessed the mother/newborn and decided to admit for further investigations (e.g. blood culture, full blood count) and management according to clinical judgment (18). In the event of death, a serious adverse event report was written by the trial clinicians based on their assessment of the cause of death, using clinical notes and investigation results.

### Outcomes and variables of interest

The primary outcome of this secondary analysis was the prevalence of IRA defined as a 1-min Apgar score of <7, based on the ICD-10 version 2019 definition (20). We chose 1-minute Apgar score instead of 5-minute Apgar score in recognition that the 1-minute score reflects the intrapartum fetal health (11, 12) and we lacked detailed data about neonatal resuscitation efforts, hence 5-minute scores were less reliable. We excluded neonates with an Apgar score of 7 as this is considered clinically reassuring and inconsistent with IRA (11, 21). We defined severe IRA as a 1-min Apgar score between 0 and 3, and mild/moderate IRA as a 1-min Apgar score between 4 and 6 (20). To explore factors associated with IRA, we developed an original conceptual framework based on a literature review and consideration of clinically and biologically plausible pathways of IRA in low-resource settings (Figure 1) (22). We differentiated the variables associated with IRA into two groups according to directionality: 1) Potential risk factors that may have contributed to the development of IRA (e.g., maternal, or obstetric complications); and 2) Factors that may have occurred due to the suspicion or presence of IRA (e.g., delivery location). The conceptual framework (Figure 1) guided our choice of variables for the univariate (unadjusted) regression analysis and informed the adjusted analysis (see details below in Statistical analysis).

**Figure 1:**
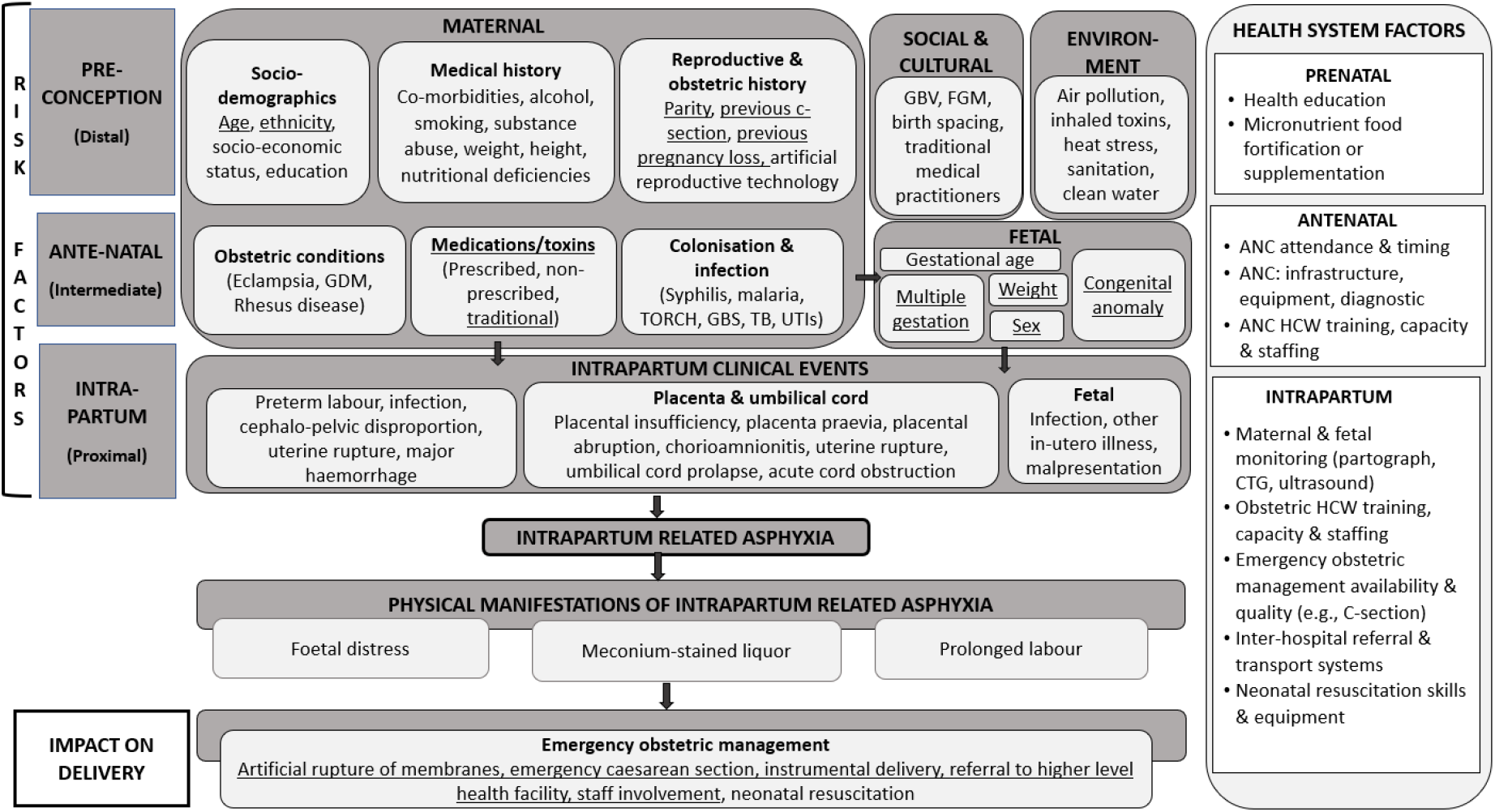
Conceptual framework for factors associated with intrapartum-related asphyxia. *(Adapted from “Perinatal risk factors for neonatal encephalopathy: anunmatched case-control study* | *ADC Foetal and Neonatal Edition)* Underlined = data available in the PregnAnZI-2 cohort ANC=Ante Natal Care; C-section=Caesarean section; CTG=Cardiotocography; FGM=Female Genital Mutilation; GBS=Group B Streptococcus; GBV= Gender-Based Violence; GDM=Gestational diabetes mellitus; HCW=Health Care Workers; TORCH= congenital Toxoplasmosis, Others (Syphilis, Hepatitis B), Rubella, Cytomegalovirus (CMV), and Herpes simplex; TB=Tuberculosis; UTIs=Urinary Tract Infections

### Operational definitions

“Women with low-risk deliveries” refers to pregnant women in low-resource health systems such as The Gambia, where antenatal detection of high-risk women/neonates is challenging; this population is representative of pregnant women thought to be at low risk at the time of enrolment. Our study was embedded into a clinical trial that excluded “known” high-risk pregnancies (acute/chronic condition, planned caesarean section, severe congenital malformation, foetal macrosomia, etc.); in practice, often high-risk pregnancies were undiagnosed and were therefore included in the trial due to the lack of rigorous antenatal diagnosis in the trial health facilities. Participants with a history of previous caesarean section or who had undergone emergency caesarean section after enrolment were not excluded from the trial; only elective caesarean section of the current pregnancy was an exclusion criterion. Also, known major congenital malformations were an exclusion criterion. However, due to limited routine antenatal detailed ultrasonography at the study health facilities,congenital malformations were frequently not antenatally diagnosed and, therefore, were included in the trial.

## Data management and statistical analysis

Relevant data was extracted from the PregnAnZI-2 trial electronic database and all analyses were done using R (version 4.1.2). Descriptive analyses were performed to determine prevalence estimates for all independent variables and adverse outcomes for the total cohort with stratification according to IRA status (low versus normal 1-minute Apgar score). Considering the complexity and multifactorial nature of IRA, with diverse factors involved at different time points (pre-conception, antepartum, and intrapartum periods), an adjusted logistic regression model was developed for each independent variable found to have p-value <0.2 for any of the categorical values on unadjusted regression. To avoid over-adjusting for multiple variables on the causal pathway (23), each model only included variables occurring before or at the same timepoint as the independent variable of interest (Figure 1). We included events in previous pregnancies (i.e. stillbirths and miscarriages) in the risk factors analysis with the assumption that the underlying causes (e.g., infectious, genetic, maternal pre-existing morbidity) of these outcomes may be recurrent or persist during the current pregnancy (24, 25) with a potential risk for IRA.

Statistical significance for the adjusted analysis was pre-determined at p-value <0.05.

Less than 1% of individuals were missing data in any variable and an available case analysis was done, i.e., individuals missing exposure or confounder variables were excluded from the adjusted analysis.

## Results

Among 6758 live births included in the analysis (Fig.2), 3.8% (257/6758) experienced IRA, with 79.0% (203/257) being mild to moderate IRA and 21.0% (54/257) severe IRA (Figure 2). Details of baseline characteristics of the study neonates and their mothers are shown in Table 1. Overall, 11.7% (30/257) and 4.7% (11/257) of newborns with IRA had low birth weight and macrosomia, respectively; and 3.1% (8/257) of newborns diagnosed with IRA had a congenital malformation.

**Table 1:** Maternal and neonatal risk factors for intrapartum-related asphyxia among low-risk pregnancies in peri-urban Gambia from 2017 to 2021.

|  | Total<br>(n=6758) | Unadjusted univariate analysis |  |  |  | Adjusted analysis |  |
| --- | --- | --- | --- | --- | --- | --- | --- |
|  |  | IRA<br>(n=257) n (%) | No IRA<br>(n=6501) n (%) | cOR (95%CI) | p-value | aOR (95% CI) | p-value |
| Maternal factors |  |  |  |  |  |  |  |
| Age (Years) |  |  |  |  | 0.42 |  | 0.99<sup>a</sup> |
| Median (IQR) | 27 (23-31) |  |  |  |  |  |  |
| 20-35 | 5459 (80.8) | 203 (3.7) | 5256 (96.3) | 1 |  | 1 |  |
| 16-19 | 631 (9.3) | 30 (4.8) | 601 (95.2) | 1.29 (0.86-1.88) |  | 1.01 (0.67-1.58) |  |
| >35 | 668 (9.9) | 24 (3.6) | 644 (96.4) | 0.96 (0.61-1.45) |  | 0.98 (0.63-1.61) |  |
| Mothers' Ethnicity |  |  |  |  | 0.41 |  |  |
| Mandinka | 2701 (40.0) | 115 (4.3) | 2586 (95.7) | 1 |  |  |  |
| Fula | 1213 (18.0) | 36 (3.0) | 1177 (97.0) | 0.69 (0.46-0.99) |  |  |  |
| Wolof | 1040 (15.4) | 39 (3.8) | 1001 (96.2) | 0.88 (0.60-1.26) |  |  |  |
| Jola | 876 (12.9) | 31 (3.5) | 845 (96.5) | 0.82 (0.54-1.22) |  |  |  |
| Others | 928 (13.7) | 36 (3.9) | 892 (96.1) | 0.90 (0.61-1.31) |  |  |  |
| Fathers' Ethnicity |  |  |  |  | 0.29 |  |  |
| Mandinka | 2612 (38.7) | 112 (4.3) | 2500 (95.7) | 1 |  |  |  |
| Fula | 1159 (17.2) | 34 (2.9) | 1125 (97.1) | 0.67 (0.45-0.99) |  |  |  |
| Wolof | 1165 (17.2) | 40 (3.4) | 1125 (96.6) | 0.79 (0.54-1.14) |  |  |  |
| Jola | 834 (12.3) | 35 (4.2) | 799 (95.8) | 0.98 (0.65-1.43) |  |  |  |
| Others | 987 (14.6) | 36 (3.6) | 951 (96.4) | 0.84 (0.57-1.23) |  |  |  |
| Parity |  |  |  |  | <0.001 |  | <0.001<sup>a</sup> |
| Para 2 | 1392 (20.6) | 50 (3.6) | 1342 (96.4) | 1 |  | 1 |  |
| Para >=4 | 2519 (37.3) | 88 (3.5) | 2431 (96.5) | 0.97 (0.68-1.39) |  | 0.80 (0.54-1.18) |  |
| Primiparous | 1681 (24.9) | 91 (5.4) | 1590 (94.6) | 1.54 (1.08-2.20) |  | 1.69 (1.16-2.46) |  |
| Para 3 | 1166 (17.2) | 28 (2.4) | 1138 (97.6) | 0.66 (0.41-1.05) |  | 0.64 (0.39-1.03) |  |
| Season of delivery<sup>b</sup> |  |  |  |  | 0.50 |  |  |
| Dry | 4575 (67.7) | 169 (3.7) | 4406 (96.3) | 1 |  |  |  |
| Rainy | 2183 (32.3) | 88 (4.0) | 2095 (96.0) | 1.09 (0.83-1.43) |  |  |  |
| *Previous stillbirth<sup>c</sup>* |  |  |  |  | <0.001 |  | 0.003<sup>a</sup> |
| No | 6642 (98.3) | 245 (3.7) | 6397 (96.3) | 1 |  | 1 |  |
| Yes | 115 (1.7) | 12 (10.4) | 103 (89.6) | 3.04 (1.50-5.64) |  | 2.79 (1.41-5.52) |  |
| *Previous caesarean section* |  |  |  |  | 0.06 |  | 0.03<sup>a</sup> |
| No | 6664 (98.6) | 250 (3.8) | 6414 (96.2) | 1 |  | 1 |  |
| Yes | 94 (1.4) | 7 (7.4) | 87 (92.6) | 2.06 (0.80-4.50) |  | 2.48 (1.12-5.53) |  |
| *Previous miscarriage<sup>d</sup>* |  |  |  |  | 0.09 |  | 0.02<sup>a</sup> |
| No | 6143 (90.9) | 226 (3.7) | 5917 (96.3) | 1 |  | 1 |  |
| Yes | 613 (9.1) | 31 (5.1) | 582 (94.9) | 1.39 (0.92-2.06) |  | 1.65 (1.08-2.52) |  |
| *Previous intake of traditional medicine<sup>e</sup>* |  |  |  |  | 0.30 |  |  |
| No | 5577 (82.6) | 206 (3.7) | 5371 (96.3) | 1 |  |  |  |
| Yes | 1177 (17.4) | 51 (4.3) | 1126 (95.7) | 1.18 (0.85-1.62) |  |  |  |
| *Pre labour rupture of membrane<sup>f</sup>* |  |  |  |  | 0.20 |  |  |
| No | 6600 (97.7) | 248 (3.8) | 6352 (96.27) | 1 |  |  |  |
| Yes | 157 (2.3) | 9 (5.7) | 148 (94.3) | 1.56 (0.69-3.09) |  |  |  |
| **Neonatal factors** |  |  |  |  |  |  |  |
| *Sex* |  |  |  |  | 0.001 |  | 0.002<sup>g</sup> |
| Female | 3262 (48.3) | 99 (3.0) | 3163 (97.0) | 1 |  | 1 |  |
| Male | 3496 (51.7) | 158 (4.5) | 3338 (95.5) | 1.51 (1.16-1.97) |  | 1.51 (1.16-1.97) |  |
| *Birth weight (kg)<sup>h</sup>* |  |  |  |  | <0.001 |  | <0.001<sup>g</sup> |
| Median (IQR) | 3.1 (2.8-3.4) |  |  |  |  |  |  |
| Normal (2.5-4.0) | 6085 (90.0) | 215 (3.5) | 5870 (96.6) | 1 |  | 1 |  |
| Low (<2.5) | 541 (8.0) | 30 (5.5) | 511 (94.5) | 1.60 (1.06-2.34) |  | 1.36 (0.88-2.08) |  |
| Macrosomia (>4.0) | 132 (2.0) | 12 (9.1) | 120 (90.9) | 2.73 (1.41-4.82) |  | 3.31 (1.78-6.15) |  |
| *Plurality of birth* |  |  |  |  | 0.09 |  | 0.06<sup>g</sup> |
| Singleton | 6580 (97.4) | 246 (3.7) | 6334 (96.3) | 1 |  | 1 |  |
| Twin | 178 (2.6) | 11 (6.2) | 167 (93.8) | 1.70 (0.82-3.17) |  | 1.90 (0.98-3.70) |  |
| *Congenital malformation<sup>i</sup>* |  |  |  |  | <0.0001 |  | <0.001<sup>g</sup> |
| No | 6716 (99.4) | 229 (3.5) | 6366 (96.5) | 1 |  | 1 |  |
| Yes | 142 (0.6) | 14 (9.9) | 128 (90.1) | 3.04 (1.59-5.39) |  | 3.01 (1.69-5.35) |  |

aOR = Adjusted odds ratio; cOR = Crude odds ratio; IQR = Interquartile range;
a) Model 1 included age of mother, fathers ethnicity, mothers ethnicity, parity, previous stillbirth, previous miscarriage, and previous caesarean section; b) Dry season between November and May; c) Previous stillbirth missing in n=1; d) Previous miscarriage missing in n=2; e) Previous intake of traditional medicine is any traditional medicine taken during the pregnancy irrespective of the route (oral, topical, or other). Information on the exact period of use or type of medicine was not recorded, missing in n=4; f) Pre-labour rupture of membranes missing in n=1; g) Model 2 included neonatal sex, birth weight, plurality of birth, and congenital malformation; h) Birth weight missing in n=2; i) Congenital malformations (minor and major) were diagnosed by trial clinicians at birth.

**Figure 2:**
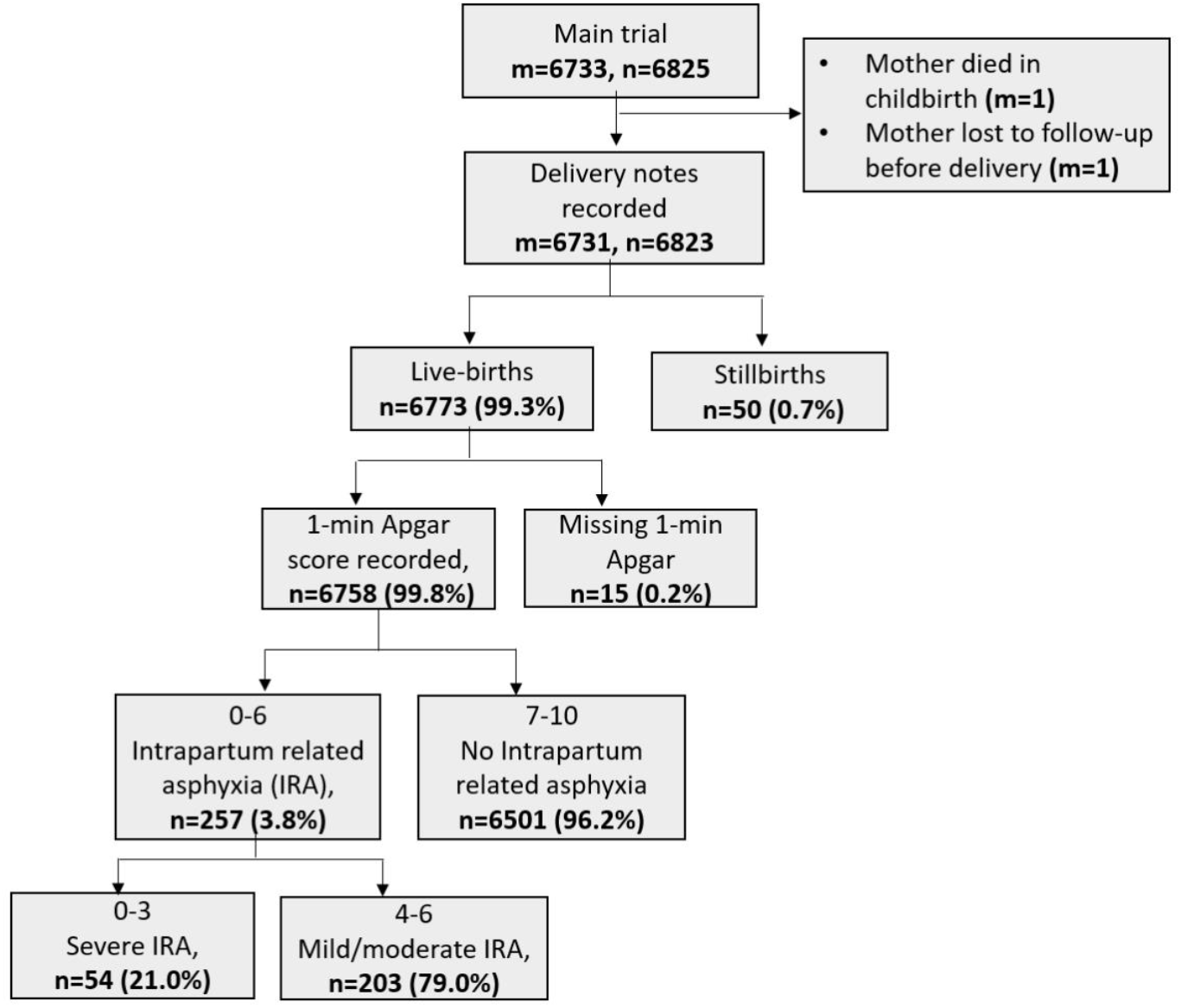
Flow chart for participants’ selection process.

### Factors associated with IRA

In the adjusted regression analysis, maternal risk factors for IRA included history of previous stillbirth (aOR: 2.79, 95%CI, 1.41-5.52, p-value=0.003), previous caesarean section (aOR: 2.48, 95%CI, 1.12-5.53, p-value=0.03) primiparity (aOR: 1.69, 95%CI, 1.16-2.46, p-value<0.001), and previous miscarriage (aOR: 1.65, 95%CI, 1.08-2.52, p-value=0.02) (Table 1). Neonatal risk factors were macrosomia (aOR: 3.31, 95%CI, 1.78-6.15, p-value<0.001),congenital malformations (aOR: 3.01, 95% CI, 1.69–5.35, p-value<0.001), and male sex (aOR: 1.51, 95% CI, 1.16-1.97, p-value=0.002) (Table 1).

Several factors that may have occurred due to clinical suspicion of prolonged/difficult delivery or foetal distress were associated with IRA. Those with the highest ORs were delivery in referral hospitals(aOR: 17.9, 95%CI, 9.36-34.10, p-value<0.001), deliveries attended by doctors (aOR: 3.25, 95%CI, 1.67-6.04, p-value<0.001) and instrumental deliveries using forceps or vacuum (aOR: 2.92, 95%CI, 1.49-5.40, p-value<0.001) (Table 2).

**Table 2:**
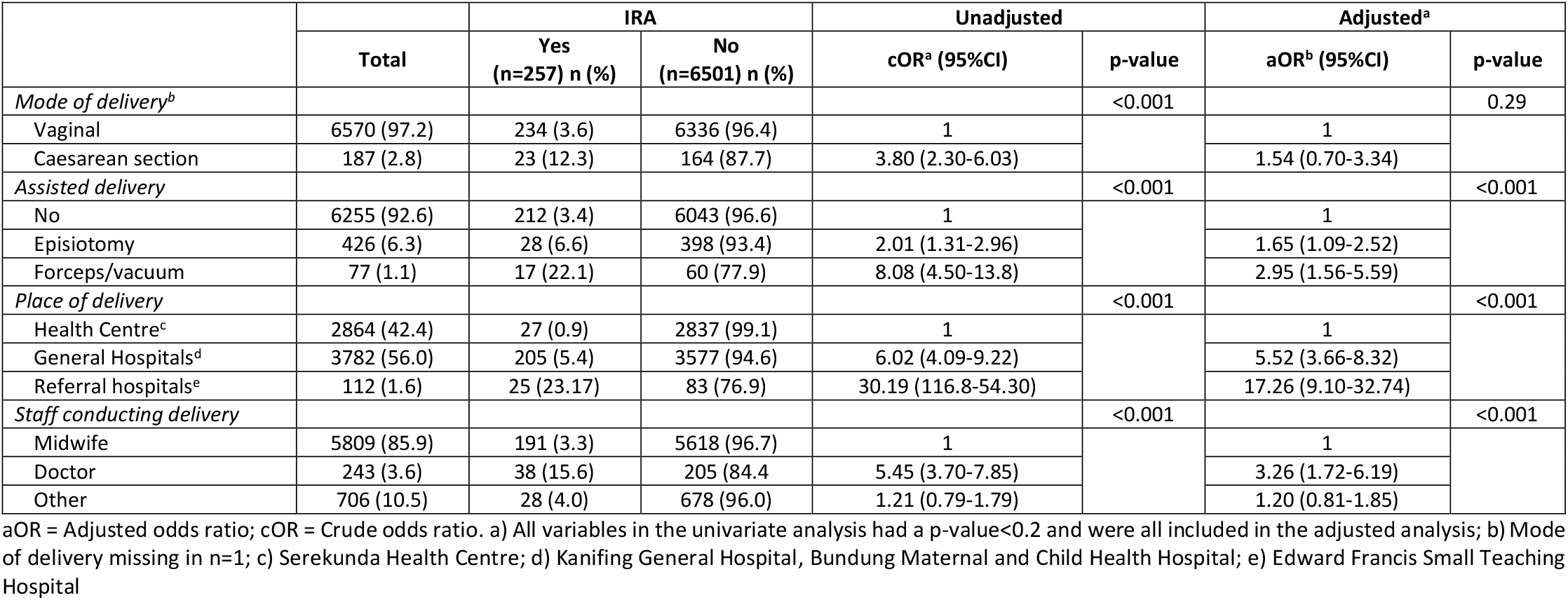
Association between intrapartum-related asphyxia and delivery circumstances among low-risk pregnancies in peri-urban Gambia from 2017 to 2021.

|  | Total | IRA |  | Unadjusted |  | Adjusted<sup>a</sup> |  |
| --- | --- | --- | --- | --- | --- | --- | --- |
|  |  | Yes<br>(n=257) n (%) | No<br>(n=6501) n (%) | cOR<sup>a</sup> (95%CI) | p-value | aOR<sup>b</sup> (95%CI) | p-value |
| *Mode of delivery<sup>b</sup>* |  |  |  |  | <0.001 |  | 0.29 |
| Vaginal | 6570 (97.2) | 234 (3.6) | 6336 (96.4) | 1 |  | 1 |  |
| Caesarean section | 187 (2.8) | 23 (12.3) | 164 (87.7) | 3.80 (2.30-6.03) |  | 1.54 (0.70-3.34) |  |
| *Assisted delivery* |  |  |  |  | <0.001 |  | <0.001 |
| No | 6255 (92.6) | 212 (3.4) | 6043 (96.6) | 1 |  | 1 |  |
| Episiotomy | 426 (6.3) | 28 (6.6) | 398 (93.4) | 2.01 (1.31-2.96) |  | 1.65 (1.09-2.52) |  |
| Forceps/vacuum | 77 (1.1) | 17 (22.1) | 60 (77.9) | 8.08 (4.50-13.8) |  | 2.95 (1.56-5.59) |  |
| *Place of delivery* |  |  |  |  | <0.001 |  | <0.001 |
| Health Centre<sup>c</sup> | 2864 (42.4) | 27 (0.9) | 2837 (99.1) | 1 |  | 1 |  |
| General Hospitals<sup>d</sup> | 3782 (56.0) | 205 (5.4) | 3577 (94.6) | 6.02 (4.09-9.22) |  | 5.52 (3.66-8.32) |  |
| Referral hospitals<sup>e</sup> | 112 (1.6) | 25 (23.17) | 83 (76.9) | 30.19 (116.8-54.30) |  | 17.26 (9.10-32.74) |  |
| *Staff conducting delivery* |  |  |  |  | <0.001 |  | <0.001 |
| Midwife | 5809 (85.9) | 191 (3.3) | 5618 (96.7) | 1 |  | 1 |  |
| Doctor | 243 (3.6) | 38 (15.6) | 205 (84.4) | 5.45 (3.70-7.85) |  | 3.26 (1.72-6.19) |  |
| Other | 706 (10.5) | 28 (4.0) | 678 (96.0) | 1.21 (0.79-1.79) |  | 1.20 (0.81-1.85) |  |

aOR = Adjusted odds ratio; cOR = Crude odds ratio. a) All variables in the univariate analysis had a p-value<0.2 and were all included in the adjusted analysis; b) Mode of delivery missing in n=1; c) Serekunda Health Centre; d) Kanifing General Hospital, Bundung Maternal and Child Health Hospital; e) Edward Francis Small Teaching Hospital

### Morbidity and mortality among newborns with IRA

Neonates with IRA were more likely to be hospitalised (OR: 11.91, 95% CI, 8.90-15.90, p-value<0.001) and die (OR: 19.56, 95% CI, 12.02–31.55, p-value<0.001) during the neonatal period than those without IRA (Table 3). Neonates with IRA represented 24% of all neonatal hospitalisations and 40% of all neonatal deaths (Figure 3). This increased risk for death and hospitalisation was higher in severe IRA but still quite significant among mild/moderate IRA (table 4).

**Table 3:** Association between neonatal admissions/deaths and intrapartum-related asphyxia among low-risk pregnancies in peri-urban Gambia from October 2017 to May 2021.

|  | Total | IRA<br>(n=257)<br>n (%) | No IRA (n=6501)<br>n (%) | Unadjusted analysis<br>OR (95% CI) | p-value |
| --- | --- | --- | --- | --- | --- |
| *Admission during the neonatal period* |  |  |  |  | <0.001 |
| No | 6370 (94.3) | 164 (2.6) | 6206 (97.4) | 1 |  |
| Yes | 388 (5.7) | 93 (24.0) | 295 (76.0) | 11.91 (8.90-15.90) |  |
| *Neonatal mortality within 24h after delivery* |  |  |  |  | <0.001 |
| Alive | 6733 (99.6) | 237 (3.5) | 6496 (96.5) | 1 |  |
| Died | 25 (0.4) | 20 (80.0) | 5 (20.0) | 109.25 (34.34-374.00) |  |
| *Neonatal mortality between 24 hours and 28 days* |  |  |  |  | <0.001 |
| No | 6413 (99.1) | 211 (3.3) | 6202 (96.7) | 1 |  |
| Yes | 60 (0.9) | 14 (23.3) | 46 (76.7) | 8.93 (4.47-16.87) |  |
| *Overall neonatal mortality by day 28<sup>a</sup>* |  |  |  |  | <0.001 |
| No | 6413 (98.7) | 211 (3.3) | 6202 (96.7) | 1 |  |
| Yes | 85 (1.3) | 34 (40.0) | 51 (60.0) | 19.56 (12.02-31.55) |  |

a) Survival status by day 28 missing in n=12

**Table 4:** Neonatal Hospitalisations and deaths associated with intrapartum-related asphyxia stratified by IRA severity in a cohort of Gambian neonates born following low-risk pregnancies.

|  | Severe<br>(n=54)<br>n (%) | Normal<br>n (%) | OR (95%CI) | p-value | Mild and Moderate<br>(n=203)<br>n (%) | Normal<br>n (%) | OR (95%CI) | p-value |
| --- | --- | --- | --- | --- | --- | --- | --- | --- |
| *Hospitalisation* |  |  |  | <0.001 |  |  |  | <0.001 |
| No | 28<br>(0.4) | 6206<br>(99.6) |  |  | 136<br>(2.1) | 6206<br>(97.9) |  |  |
| Yes | 26<br>(8.1) | 295<br>(91.9) | 19.5<br>(10.8-35.0) |  | 67<br>(18.5) | 295<br>(81.5) | 10.35<br>(7.43-14.3) |  |
| *Mortality within 24h* |  |  |  | <0.001 |  |  |  | <0.001 |
| No | 40<br>(0.6) | 6496<br>(99.4) |  |  | 197<br>(2.9) | 6496<br>(97.1) |  |  |
| Yes | 14<br>(73.7) | 5<br>(26.3) | 448.5<br>(144-1600) |  | 6<br>(54.5) | 5<br>(45.5) | 39.5<br>(9.9-164.7) |  |
| *Mortality between 24h to 28 days* |  |  |  | <0.001 |  |  |  | <0.001 |
| No | 32<br>(0.5) | 6202<br>(99.5) |  |  | 179<br>(2.8) | 6202<br>(97.2) |  |  |
| Yes | 5<br>(9.8) | 46<br>(90.2) | 21.0<br>(6.10-57.80) |  | 9<br>(16.4) | 46<br>(83.6) | 6.8<br>(2.9-14.3) |  |
| *All mortality by day 28<sup>a</sup>* |  |  |  | <0.001 |  |  |  | <0.001 |
| No | 32<br>(0.5) | 6202<br>(99.5) |  |  | 179<br>(2.8) | 6202<br>(97.2) |  |  |
| Yes | 19<br>(27.1) | 51<br>(72.9) | 71.8<br>(36.0-140.4) |  | 15<br>(22.7) | 51<br>(77.3) | 10.2<br>(5.2-18.8) |  |

a) survival status missing in n=3 and n=9 in severe and mild/moderate IRA respectively.

**Figure 3:**
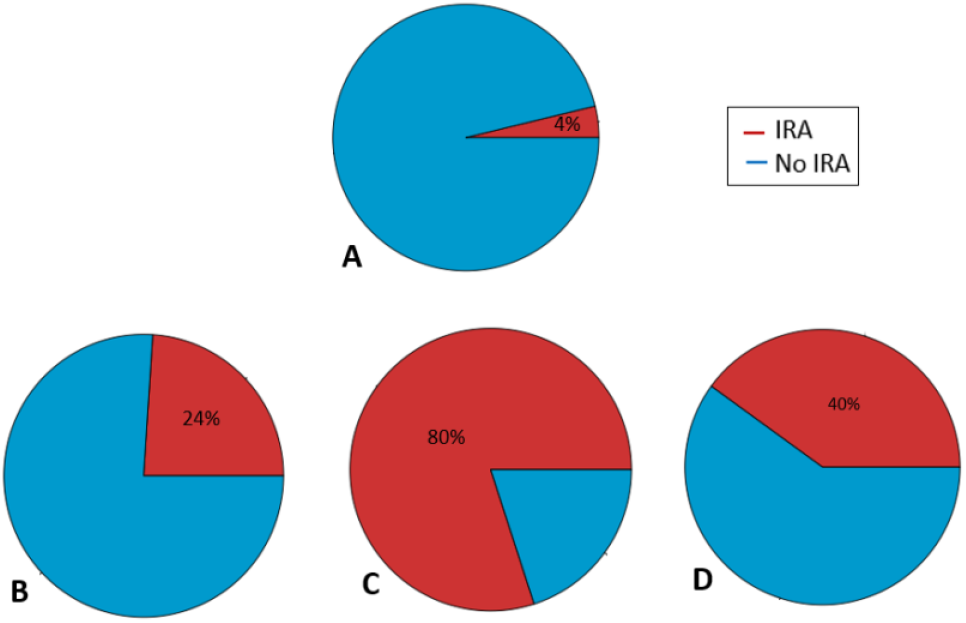
Prevalence of neonates with intrapartum-related asphyxia (IRA) at birth versus prevalence of neonates with IRA among neonatal admissions and deaths(A) Prevalence of neonates with IRA at birth; (B) Prevalence of neonates with IRA among admissions (0-28 days); (C) Prevalence of neonates with IRA among neonatal deaths (0-24H after birth); (D) Prevalence of neonates with IRA among overall neonatal deaths (0-18 days

## Discussion

IRA is a major cause of neonatal morbidity and mortality among neonates born in peri-urban health facilities in The Gambia, even for low-risk pregnancies. Although less than 4% (1 in 25) of neonates experienced IRA, these neonates represented 24% of all neonatal hospitalisations and 40% of all neonatal deaths in our cohort. We identified an important modifiable neonatal risk factor for IRA (i.e., macrosomia). In addition, we detected several maternal risk factors such as primiparity, history of previous stillbirth, or previous miscarriage that could signal a high-risk phenotype and trigger enhanced antenatal surveillance and early intervention to prevent IRA.

The prevalence of IRA in our cohort was lower than reported in other African studies, such as in Ethiopia (19.3%) (7) and Nigeria (29.4%) (26). However, those studies were conducted at referral hospitals where high-risk deliveries, including caesarean sections, were more common, possibly explaining the higher prevalence of IRA. We also excluded women with known acute or chronic conditions (27) and, by selecting out women with detectable risk of intrapartum complications, this may have under-estimated the prevalence of IRA.

In our study, IRA was recorded in 2 out of 5 neonatal deaths, and 4 out of 5 neonatal deaths within 24hours of delivery. Therefore, the contribution of IRA to neonatal mortality is at least two-to three-fold higher than shown by previous studies in West Africa (such as Nigeria, 15.7%, and Ghana, 21.8%), despite the prevalence of IRA among neonates being higher (28, 29). A possible explanation for the higher mortality in our setting compared to others in the sub-region is the lack of specialised care at delivery, both in terms of infrastructure and human resources. Indeed, in Nigeria, paediatricians and obstetricians were 24h available for all high-risk deliveries (28). In Ghana, paediatricians were present at all deliveries in which IRA was suspected, and all neonates at high-risk of neonatal encephalopathy were transferred to the Neonatal Intensive Care Unit, regardless of their Apgar scores (29). This contrasts with our setting where trained midwives and nurses in the delivery room, medical officers were always available, but specialists (paediatricians and obstetricians) were inconsistently present due to their low number. As a result, delays in appropriate management of maternal and neonatal complications were occasionally observed. The risk for poor outcomes observed among neonates with IRA in our study was higher within the severe cases as expected but still significant among neonates with mild/moderate cases suggesting a possible subjectivity of the Apgar scoring and probable misclassification. Previous studies showed the same results (30). This highlights that prompt and effective resuscitation and post-resuscitation management are critical to reducing mortality among neonates with IRA even in mild/moderate cases (31).

We identified both maternal and neonatal factors associated with IRA. From the maternal perspective, primiparity was an important risk factor for IRA. In line with previous studies in Nigeria (28) and Uganda (32), primiparous women often have prolonged labour, and are likely to have cephalo-pelvic disproportion, increasing the risk of obstructed labour (33). Mothers with a history of previous stillbirth or miscarriage were also at higher risk of delivering a neonate with IRA. This clustering effect may be explained by either maternal biomedical factors (such as cephalo-pelvic disproportion), or biological factors (such as maternal chronic or acute conditions including infection, eclampsia, diabetes) which have been previously described in low- and middle-income countries (34–36). From the neonatal perspective, male gender was associated with IRA; as previously described in Togo (37) and Ethiopia (38). Although the mechanisms underlying this sex difference are not fully understood, females seem to have better in-utero hypoxic tolerance compared to male neonates (39–41). Macrosomic neonates were also at higher risk of IRA, as previously described in studies from the region (42, 43). Macrosomic neonates often present with peripartum complications such as obstructed second stage of labour (44, 45) or shoulder dystocia, highlighting the importance of foetal growth monitoring and planned Caesarean-section to mitigate this risk. Congenital malformation is a risk factor for IRA, as shown also in Tanzania (46) and Nigeria (47). Although the exact causal pathway is not fully understood, a study in Finland found an association between congenital malformations and breech delivery (48). It is well known that foetal malposition, especially breech presentation is associated with an increased risk of hypoxic-anoxic events (49) which may progress into IRA (50). Early detection of foetal malposition by routine detailed sonography would enable planning for safe delivery. (51, 52).

Neonates with IRA were more likely to have been delivered in general and referral teaching hospitals, and were more frequent among deliveries conducted by doctors, consistent with studies conducted in Uganda (53) and Ethiopia (54). In addition, neonates with IRA were more frequent among instrumental delivery (forceps or vacuum), similar to an Ethiopian study (55). Reverse causality is the most likely explanation for these results. Indeed, complicated deliveries with high risk of IRA are indications for instrumental deliveries and these women are referred to highly equipped health facilities with more qualified staff such as midwives and medical doctors. These findings indicate that health workers were able to detect deliveries affected by IRA and make appropriate referrals and management decisions. However, these actions did not translate into improved neonatal outcomes as shown by the high mortality of neonates with IRA, reflecting the importance of timely decision-making, with reasons for delayed referrals or obstetric interventions likely to be multifactorial (56). More research is required to understand the barriers and enablers for timely and optimal management of high-risk deliveries in varying West African settings to prevent avoidable neonatal deaths.

A limitation of this study is the potential for large intra- and inter-observer variability in Apgar score assignation, despite targeted training of health workers. The observed association between IRA and mortality indicates that specificity is probably high despite the potential for subjectivity. Secondly, overall neonatal mortality in our cohort is probably an underestimation of neonatal mortality due to both the healthy effect of research participation (57) and the exclusion of women with acute or chronic co-morbidities. The healthy effect of trial participation should have impacted all endpoints; therefore, decreasing the overall mortality rate but not necessarily the distribution of causes or risk factors. In summary, our findings are generalisable to a population of low-risk pregnant women delivering in low-resource West African hospital settings where antenatal detection of high-risk women and neonates is challenging yet may not apply to other populations or settings with more comprehensive antenatal diagnostics.

## Conclusions

Our study highlights the high prevalence of IRA among neonatal deaths, even in a low-risk West African population, and several risk factors for IRA. Attention to modifiable risk factor (i.e., macrosomia) is required to decrease the prevalence of IRA. Better control of maternal diabetes and identification of foetal macrosomia for safe delivery plan are needed to modify the association of macrosomia with IRA. In addition, effective interventions to improve antenatal and obstetric care and post-delivery management of neonates exposed to IRA, such as prompt resuscitation by trained, skilled staff, are crucial if global neonatal mortality reduction targets are to be met by 2030.

## Data Availability

Data may be obtained from a third party and are not publicly available. The clinical data has been collected following provision of informed consent under the prerequisite of strict participant confidentiality. Qualified researchers may request access with the Gambia Government and MRC Joint Ethics Committee. The review process and release of data will be facilitated by MRC Unit The Gambia (http://www.mrc.gm/) through the Head of Governance at MRCG. Access will not be unduly restricted.

## List of abbreviations

IRA: Intrapartum-Related Asphyxia
DALY: Disability Adjusted Life Years
SSA: Sub-Saharan Africa
SDG: Sustainable Development Goal
SHC: Serekunda Health Centre
BMCHH: Bundung Maternal and Child Health Hospital
eCRF: electronic Case Report Form

## DECLARATIONS

### Ethics approval and consent to participate

The PregnAnZI2 trial was approved by The Gambia Government/Medical Research Council Unit The Gambia (MRCG) Joint Ethics Committee, the Comité d’Ethique pour la Recherche en Santé (CERS) and the Ministry of Health of Burkina Faso, and the London School of Hygiene and Tropical Medicine (LSHTM) Ethics Committee. This included approval for further research utilising trial data. During antenatal care visits all participants provided written informed consent to participate in the trial and were free to withdraw at any time. They also consented for the use of their data in further research.

### Availability of data and materials

Data may be obtained from a third party and are not publicly available. The clinical data has been collected following provision of informed consent under the prerequisite of strict participant confidentiality. Qualified researchers may request access with the Gambia Government/MRC Joint Ethics Committee. The review process and release of data will be facilitated by MRC Unit The Gambia (http://www.mrc.gm/) through the Head of Governance at MRCG. Access will not be unduly restricted.

## Authors’ contributions

AR and UdA conceived and designed the initial trial. HT contributed to the trial design and implementation. NB designed this study with input from AR, HB, and CB. NB performed the analysis with full access to the data and input from CB. NB drafted the initial manuscript with input from HB, AR, CB, FS, and UdA. AR gave oversight to the work as guarantor and accepts full responsibility for finished work and controlled the decision to publish. All co-authors gave their approval for the publication of the final version of the manuscript.

### Consent to publish

Not applicable

## Acknowledgements

We thank the project managers, Asheme Mahmoud and Bakary Fatty and all the field, data-management, and laboratory teams for providing support to conduct the main trial. We also thank the BMCHH and SHC leadership board and staff for their support. We are grateful to all the mothers and their neonates who participated to this study.

